# Periodontitis and Associated Factors Among Dental Outpatients at Tertiary Hospitals in Dhaka

**DOI:** 10.64898/2026.11.03.26367143

**Authors:** Sadia Mehjabeen Kanta, Fahmida Faizha Shama, Md. Golam Sharower, Sifat Sharmin, Hafiza Sultana

**Affiliations:** Department of Public Health, Canadian University of Bangladesh; National Institute of Preventive and Social Medicine, Bangladesh; International Network Doctor’s Journal, Bangladesh

**Keywords:** Periodontitis, Oral hygiene, Tobacco use, Diabetes mellitus

## Abstract

**Objective:** To identify the factors associated with periodontitis among dental outpatient departments at tertiary hospitals in Dhaka.

**Methods:** This cross-sectional study included 227 patients from dental outpatient departments of three tertiary-level hospitals in Dhaka, Bangladesh by intraoral examinations and in-person interviews using a pretested semi-structured questionnaire through Simplified Oral Hygiene Index (OHI-S) and the Community Periodontal Index (CPI) component of the CPITN.

**Results:** About 55% were classified as having periodontitis. Both diabetes mellitus (p < 0.05) and harmful oral habits (smoking, smokeless tobacco, and constant tooth picking) were significantly associated with periodontitis (p < 0.05). Participants with diabetes had 2.87 times higher odds of periodontitis (OR: 2.870, 95% CI: 1.516-5.441). The odds of having periodontitis were 7.27 times higher for smokeless tobacco users (OR: 7.271, 95% CI: 3.609-14.651), and smokers had 3.69-fold higher odds of periodontitis (OR: 3.687, 95% CI: 1.701-7.988). A strong positive correlation was observed between poor oral hygiene status and the presence of periodontitis (r = 0.947, p < 0.001).

**Conclusion:** Diabetes, smoking and poor oral hygiene are key determinants of periodontitis which underscore the need for targeted oral health promotion, regular screening and integration of periodontal care into geriatric services.

## 1 Introduction

Periodontitis is a prevalent chronic inflammatory disease characterised by progressive destruction of the supporting structures of the teeth, including the periodontal ligament and alveolar bone, ultimately leading to tooth loss if left untreated [1]. It is one of the most common oral diseases that affects older people worldwide. It is often linked to a decline in oral hygiene, systemic health conditions, and a lower quality of life [2]. Older adults with poor oral health may experience issues, including tooth decay, gum disease, dry mouth, tooth loss, and even oral precancerous or cancerous lesions [2,3].

Physiological ageing makes older adults susceptible to periodontal diseases, one of the most common chronic conditions [4]. Studies conducted in both the Bangladeshi and global contexts have shown that older people frequently have serious unmet oral health needs and poor oral hygiene habits [3,5]. Older populations need to receive effective oral health care to ensure quality of life, functional performance, and overall systemic health[4].

Several sociocultural and behavioural risk factors, including the consumption of betel quid and tobacco, prevalent in South Asia, have been significantly associated with periodontal tissue damage, intensifying gingival inflammation and attachment loss [6,7]. Both smoking and the use of smokeless tobacco are acknowledged as significant factors in periodontal disease progression and severity [7]. Moreover, factors such as inadequate oral hygiene, ageing, genetic predisposition, certain medications, and psychological stress have been associated with the aetiology of periodontal disease [8–10]. Proper oral hygiene and preventive care are essential for preserving periodontal health, especially in older adults, who may encounter additional challenges such as diminished mobility, lack of awareness, or limited access to dental services [9]. Hyperglycemia compromises immune function and stimulates inflammatory responses, consequently heightening the susceptibility and severity of periodontal disease in individuals with diabetes mellitus, which exhibits a well-established bidirectional relationship with periodontitis [1,11].

With the increasing proportion of elderly people in Bangladesh and globally, it is important to meet their oral health needs to improve their overall well-being. Identifying modifiable behavioural and clinical factors associated with periodontitis will help inform targeted oral health promotion and preventive strategies. Therefore, this study aimed to identify factors associated with periodontitis among older adults attending dental outpatient departments in Dhaka, Bangladesh.

## 2 Materials & methods

### 2.1 Study Design and Setting

A descriptive cross-sectional study was conducted from January to December 2022 involving older adults visiting the dental outpatient departments of three tertiary hospitals in Dhaka, Bangladesh: Shaheed Suhrawardy Medical College & Hospital, Dhaka Dental College Hospital, and Probin Hospital, Agargaon.

The present study was conducted using data from the same parent study population as a previously published preprint. Although both studies were derived from the same dataset and share participant sociodemographic characteristics, they address distinct research objectives and examine different outcomes and analyses [12].

### 2.2 Study Population and Sampling

We recruited Older Adults aged 46 years or older in the study by convenience sampling. Individuals who had edentulism, less than two teeth in each sextant, or physical/mental disabilities were excluded. A total of 227 participants were included in the analysis. The sample size was calculated based on an expected proportion of periodontitis of 82% among the elderly, obtained from the literature search [13], with a 95% confidence level and 5% margin of error.

The inclusion criteria for participants were older adults aged 46 years and above attending the dental OPD who were willing to participate and provide informed written consent. The exclusion criteria were patients with mental or physical disabilities, those who were edentulous, and those who did not have at least two teeth in each sextant.

### 2.3 Data Collection

Written informed consent was obtained before participation. Data were collected through face-to-face interviews using a pretested semi-structured questionnaire, followed by clinical oral examinations performed under adequate illumination.

Oral hygiene status was assessed using the Simplified Oral Hygiene Index (OHI-S). Periodontal health status was evaluated using the Community Periodontal Index (CPI) component of the Community Periodontal Index of Treatment Needs (CPITN). Participants were categorised as having periodontitis (CPI ≥3) or non-periodontitis (CPI ≤2), in line with a previous study [13]. Clinical examination of plaque and calculus was performed using a dental explorer and mouth mirror, while periodontal pocket depth was measured with a WHO CPI probe.

Key behavioural and clinical variables were defined as follows: smoking and smokeless tobacco use were self-reported; diabetes mellitus was defined as self-reported physician-diagnosed diabetes; harmful oral habits included habitual tooth picking and clenching or grinding teeth.

### 2.4 Statistical Analysis

Data were analysed using SPSS version 26. Descriptive statistics (frequencies, percentages, and means) were used for sociodemographic and clinical variables. We used Inferential statistics, including chi-square and Fisher’s exact tests, and Pearson’s correlation test, to assess the association and linear relationship between variables. A p-value <0.05 was considered statistically significant. Binary Logistic regression analysis was performed to predict the potential factors, adjusting for relevant confounders to estimate both unadjusted and adjusted odds ratios with 95% confidence intervals.

## 3 Results

A total of 227 patients (mean age: 55.8 ± 8.2 years) participated in the study. Of these, 51.1% were female, and 48.9% were male. Nearly half (48.5%) were in the 46–55-year age group, while only 1.8% were aged >75 years. Detailed characteristics of the study population have been reported previously[12].

Tobacco use was common: 24.2% of participants smoked, and 35.2% used smokeless tobacco. Alcohol consumption (1.3%), habitual tooth-picking (3.1%), and habit of clenching or grinding teeth (0.9%) were rare (Table 1).

**Table 1.**
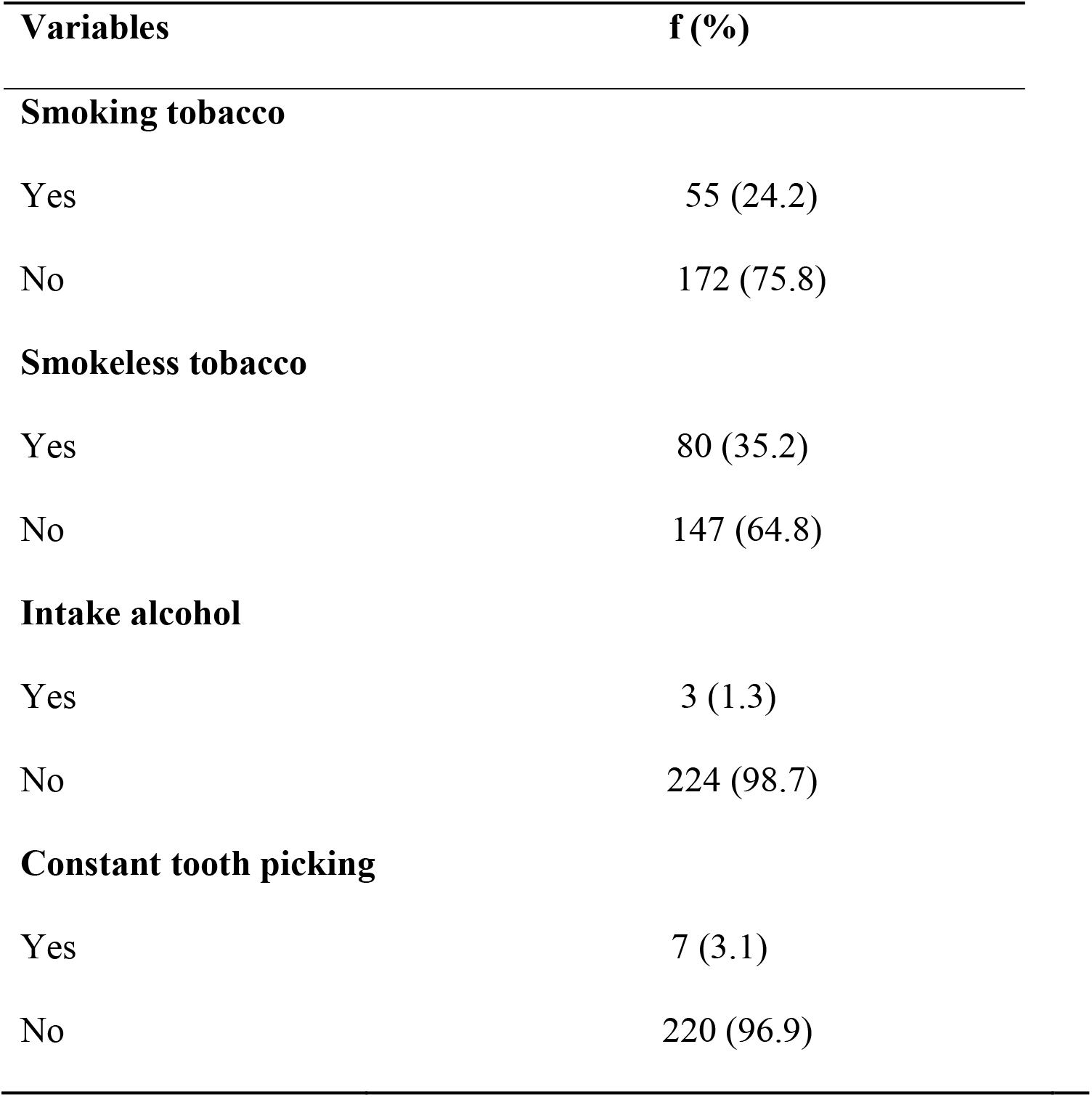
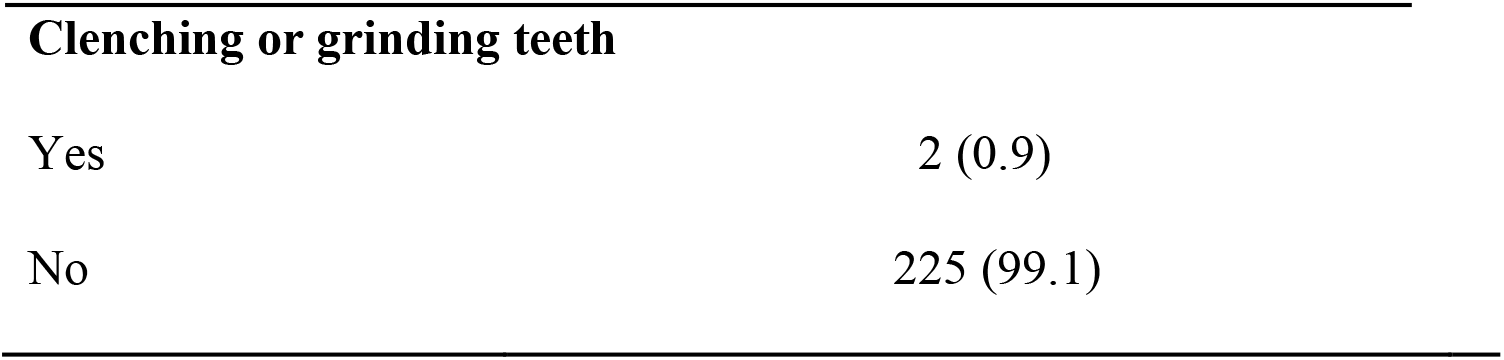
Distribution of participants by deleterious oral habits.

The study found that 31 (13.66%) had no signs of periodontal disease. A small proportion, 17 (7.49%), exhibited bleeding on probing. Calculus with plaque detectable by probing was observed in 53 individuals (23.35%). Additionally, 62 participants (27.31%) had pathological pockets measuring 4–5 mm, while 64 (28.19%) had pockets of 6 mm or more (Table 2).

**Table 2.** Periodontal Health Status of Study Participants.

| **Community Periodontal Index** | **f (%)** |
|---|---|
| No periodontal disease | 31(13.66) |
| Bleeding on probing | 17 (7.49 |
| Calculus with plaque felt by probing | 53 (23.35) |
| Pathological pocket 4-5 mm | 62 (27.31) |
| Pathological pocket 6mm or more | 64(28.19) |

Figure 1 depicts the Distribution of diabetes and periodontitis among participants. Overall, 55.5% of participants were classified as having periodontitis. Of the total sample, 39.2% reported a diagnosis of diabetes.

**Figure 1.**
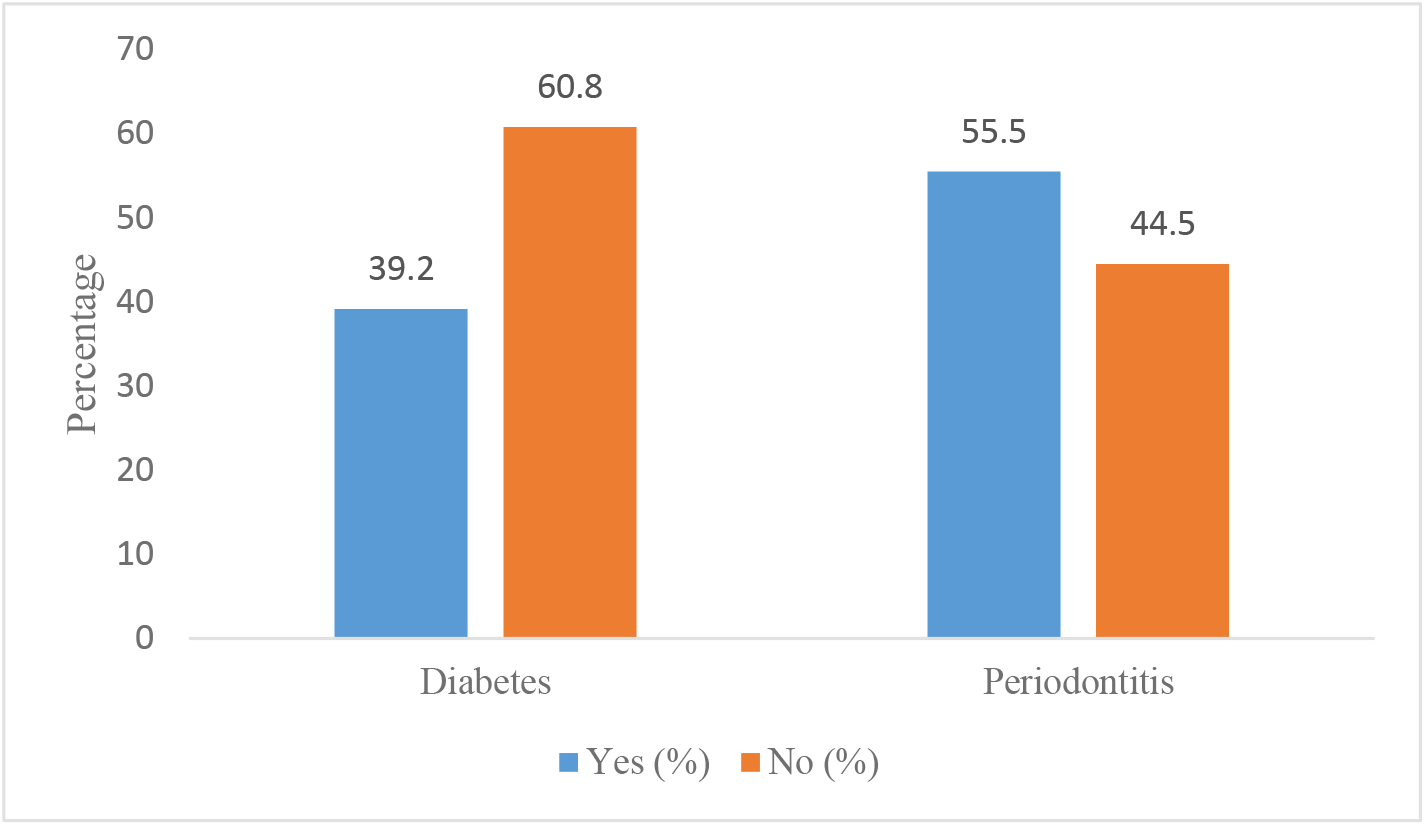
Distribution of diabetes and periodontitis among participants.

Table 3 shows the association between diabetes and periodontitis among participants. The proportion of periodontitis was significantly higher among diabetics compared with non-diabetics (71.9% vs. 44.9%; p<0.001).

**Table 3.** Association Between Diabetes and Presence of Periodontitis.

| **Diabetes** | **Presence of periodontitis** |  | **p value** |
|---|---|---|---|
|  | **Periodontitis**<br>**f (%)** | **Non-periodontitis**<br>**f (%)** |  |
| Yes | 64 (71.90%) | 25 (28.10%) | <0.001 |
| No | 62 (44.90%) | 76 (55.10%) |  |

Periodontitis was significantly associated with tobacco use. The proportion was higher among smokers (78.2% vs. 48.3%; p<0.001) and smokeless tobacco users (82.5% vs. 40.8%; p<0.001). Constant tooth-picking was also associated with a higher percentage (100% vs. 54.1%; p=0.018). Although all participants who consumed alcohol or reported bruxism had periodontitis, these associations were not statistically significant due to the small numbers (Table 4).

**Table 4.** Association between deleterious oral habits and periodontitis.

| **Variable** | **Periodontitis f (%)** | **Non-periodontitis f (%)** | **p-value** |
|---|---|---|---|
| **Smoking Tobacco** |  |  |  |
| Yes | 43 (78.20%) | 12 (21.80%) | <0.001 |
| No | 83 (48.30%) | 89 (51.70%) |  |
| **Smokeless Tobacco** |  |  |  |
| Yes | 66 (82.50%) | 14 (17.50%) | <0.001 |
| No | 60 (40.80%) | 87 (59.20%) |  |
| **Alcohol intake** |  |  |  |
| Yes | 3 (100.00%) | 0 (0.00%) | 0.25* |
| No | 123 (54.90%) | 101 (45.10%) |  |
| **Constant tooth picking** |  |  |  |
| Yes | 7 (100.00%) | 0 (0.00%) | 0.018* |
| No | 119 (54.10%) | 101 (45.90%) |  |
| **Clenching or grinding teeth** |  |  |  |
| Yes | 2 (100.00%) | 0 (0.00%) | 0.50* |
| No | 124 (55.10%) | 101 (44.90%) |  |

Note: p-values derived from the chi-square test unless otherwise indicated. \*Fisher's exact test applied.

Diabetes, smoking tobacco, and smokeless tobacco use had significant associations with periodontitis (Table 5). Participants with diabetes were 3.14 times more likely to develop periodontitis compared to those without diabetes (ORun-adj: 3.138, 95% CI: 1.773–5.555, *p* < 0.0001; ORadj: 2.870, 95% CI: 1.516–5.441, *p* = 0.001). Similarly, smokers had 3.84 times greater odds of periodontitis than non-smokers (ORun-adj: 3.842, 95% CI: 1.896–7.787, *p* < 0.0001; ORadj: 3.687, 95% CI: 1.701– 7.988, *p* = 0.001). In addition, those who use smokeless tobacco were 6.84 times more likely to develop periodontitis compared to non-users (ORun-adj: 6.836, 95% CI: 3.519–13.277, *p* < 0.0001; ORadj: 7.271, 95% CI: 3.609–14.651, *p* = 0.001).

**Table 5.**
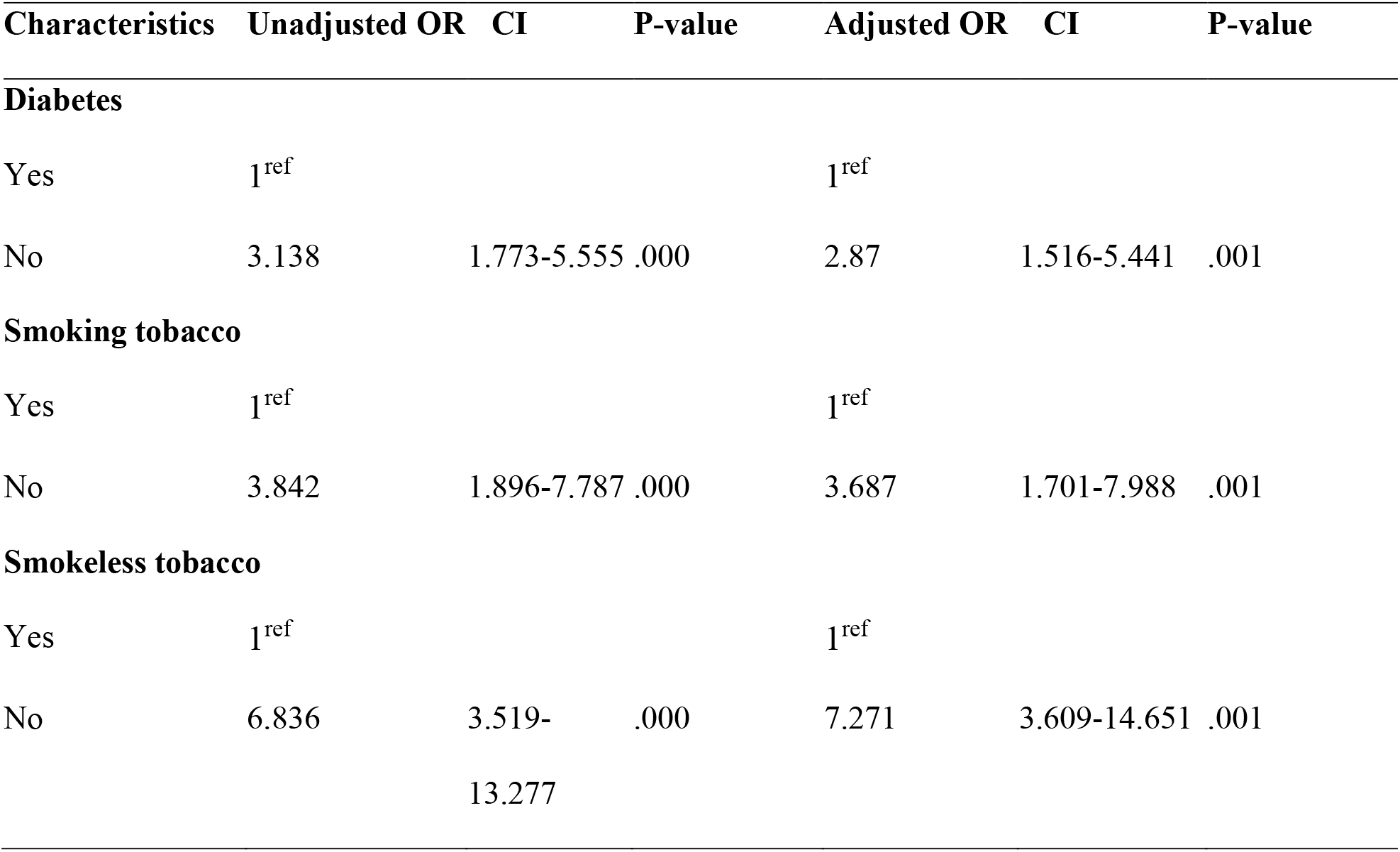
Logistic Regression Analysis of Factors Associated with Periodontitis.

| **Characteristics** | **Unadjusted OR** | **CI** | **P-value** | **Adjusted OR** | **CI** | **P-value** |
|---|---|---|---|---|---|---|
| **Diabetes** |  |  |  |  |  |  |
| Yes | 1<sup>ref</sup> |  |  | 1<sup>ref</sup> |  |  |
| No | 3.138 | 1.773-5.555 | .000 | 2.87 | 1.516-5.441 | .001 |
| **Smoking tobacco** |  |  |  |  |  |  |
| Yes | 1<sup>ref</sup> |  |  | 1<sup>ref</sup> |  |  |
| No | 3.842 | 1.896-7.787 | .000 | 3.687 | 1.701-7.988 | .001 |
| **Smokeless tobacco** |  |  |  |  |  |  |
| Yes | 1<sup>ref</sup> |  |  | 1<sup>ref</sup> |  |  |
| No | 6.836 | 3.519-13.277 | .000 | 7.271 | 3.609-14.651 | .001 |

Variables including alcohol intake, constant tooth picking, and clenching or grinding teeth were not included in the logistic regression analysis due to very small numbers of participants in some categories, which resulted in unstable odds ratio estimates.

A strong positive correlation was observed between oral hygiene status and periodontal health (Table 6). Lower OHI-S scores were strongly associated with higher CPI scores (r=0.947, p<0.001).

**Table 6.** Correlation between oral hygiene index and periodontal index.

|  |  | **Oral hygiene index** | **Periodontal index** |
|---|---|---|---|
| **Oral hygiene index** | Pearson Correlation | 1 | .947*** |
|  | p-value | - | 0.000 |
| **Periodontal index** | Pearson Correlation | .947*** | 1 |
|  | p-value | 0.000 | - |

Note: Value p < 0.05= presence of correlation (\* significant); correlation -1 <r<1 shows direction and strength of correlation.

## 4 Discussion

This study found that over half of the participants had periodontitis, with significant associations observed for age, gender, socioeconomic status, diabetes mellitus, deleterious oral habits, and oral hygiene. These results align with global and regional studies, which indicate that the proportion of periodontitis among elderly populations varies from 40% to 75%, depending on the population characteristics and diagnostic criteria [14,15]. The chronic and progressive nature of periodontal disease, along with its association with systemic health and quality of life, underscores the need for preventive measures and targeted interventions for older adults.

Among participants with diabetes, 71.9% had periodontitis, indicating a significant association between the two conditions. This result supports the established bidirectional relationship between diabetes and periodontal disease [1,16]. Chronic hyperglycemia increases the production of inflammatory cytokines, changes the metabolism of collagen, and impairs neutrophil function, all of which lead to periodontal destruction [17]. In contrast, periodontal infections worsen glycemic control by increasing insulin resistance and systemic inflammation [18]. Additionally, new research shows that diabetics’ gut microbial dysbiosis and oral microbiome may exacerbate the inflammatory environment, worsening the deterioration of periodontal tissue [19].

This study found that smoking, smokeless tobacco use, and habitual tooth picking were significantly associated with periodontitis. Tobacco consumption remains the leading determinant of periodontal disease [20,21]. Furthermore, Smoking weakens the immune system, delays the healing of wounds, and decreases the efficacy of periodontal therapies [22]. Smokeless tobacco use is most associated with gingival recession and periodontal disease [23]. Although tooth picking is often intended as an oral hygiene practice, it can cause mechanical trauma and gingival irritation when performed excessively or incorrectly, leading to periodontal pockets and loss of attachment [24].

A strong and statistically significant positive correlation was found between the periodontal index and the oral hygiene index. The proportion of periodontitis was considerably higher in older adults with poor oral hygiene, which is consistent with the study that found fair to poor oral hygiene increases the risk of periodontitis[25]. Infrequent tooth brushing was found to be associated with the severity of periodontal disease, according to a systematic review [26].

Our study demonstrated that diabetes, smoking tobacco, and smokeless tobacco use were significantly associated with periodontitis. In line with earlier research showing that hyperglycemia exacerbates the loss of periodontal tissue, older adults with diabetes had almost three times the risk of developing periodontitis compared to their non-diabetic counterparts [27]. Similarly, tobacco users had significantly higher odds of developing periodontitis, and both smoking and smokeless tobacco were found to be significant factors. According to this study, smokers had 3.84 times greater odds of periodontitis than non-smokers. Ayub et al. [28] found that the odds ratio for periodontitis among current smokers varies from 1.5 to 7.3 when compared to nonsmokers, depending on the severity of condition.

An important factor in determining tooth loss, coronal root caries, and periodontal disease is the length of tobacco use, and older adults who smoke are almost three times more likely to develop severe periodontal disease [29].

In this study, Smokeless tobacco (SLT) users had more than sevenfold increased odds, underscoring the substantial periodontal damage associated with this habit. Studies in India found that consuming smokeless tobacco led to more periodontal tissue loss [30–32]. A Systematic Review on the Association between Smokeless Tobacco Use and Risk of Periodontitis in Asian Countries also suggested that, compared to non-users, SLT users had worse periodontal health [33]. These results underline the pressing need for targeted preventive measures among senior populations and support the well-established roles of diabetes and tobacco as significant modifiable factors associated with periodontitis.

This study has several limitations. The cross-sectional design limits the ability to infer causality. The use of convenience sampling and reliance on self-reported habits may introduce selection and reporting biases. The clinical parameters were assessed only once, and radiographic examination, which could have provided more detailed insights into alveolar bone loss, was not included due to resource constraints. Furthermore, variables such as alcohol intake, constant tooth picking, and clenching or grinding teeth were not included in the logistic regression analysis due to the very small number of participants in certain categories, which limited the stability of the estimates. Although these factors may still influence periodontal health, further studies are required to elucidate their relationship with periodontitis in this population.

## 5 Conclusion

The study highlights the high proportion of periodontitis among older adults in Dhaka, which is significantly associated with age, gender, education, occupation, income, diabetes mellitus, some harmful oral habits, and poor oral hygiene. These results underline the necessity of comprehensive oral health initiatives targeting the ageing populations, especially those with chronic illnesses such as diabetes. Promoting oral health literacy and expanding access to preventive dental care could reduce the burden of periodontal disease and improve the quality of life.

## Data Availability

All data produced in the present study are available upon reasonable request to the authors

## Funding

The authors received no specific funding for this work.

## Acknowledgement

The authors would like to thank all participants in this study.

## Ethical Consent and Permission for Data Collection

This study was approved by the Institutional Review Board of the National Institute of Preventive and Social Medicine (NIPSOM), Bangladesh (Ref. No.: NIPSOM/IRB/2017/09). The Declaration of Helsinki (revised version) was followed to conduct the study. Both written and verbal consent were obtained before initiating the interview. A brief on the aims and objectives was given to the participants. Participants who agreed to provide consent were finally included in the study.

## Conflict of Interests

All authors declared no conflicts of interest.

## Data availability statement

The data that support the findings of the study are available from the corresponding author upon reasonable request.

## Notes

### Competing Interest Statement

The authors have declared no competing interest.

### Author Declarations

This study was approved by the Institutional Review Board of the National Institute of Preventive and Social Medicine, Bangladesh (Ref. No: NIPSOM/IRB/2017/09). The Declaration of Helsinki (revised version) was followed to conduct the study. Both written and verbal consent were obtained before initiating the interview. A brief on the aims and objectives was given to the participants. Participants who agreed to provide consent were finally included in the study.

